# Outcomes of Endovascular Thrombectomy in Patients with Cerebral Venous Thrombosis: A Cohort Study of 36,005 Patients

**DOI:** 10.1101/2024.09.30.24314676

**Authors:** Aryan Malhotra, Ankita Jain, Eris Spirollari, Rachid Kaddoura, Melinda Christine Arthur, Ariel Sacknovitz, Mohammed B. Nawaiseh, Chaitanya Medicherla, Gurmeen Kaur, Justin Santarelli, Chirag D. Gandhi, Fawaz Al-Mufti

## Abstract

**Introduction/Purpose:** Cerebral venous thrombosis (CVT) is a rare condition that presents significant treatment challenges and has traditionally been managed with anticoagulation. However, for patients who fail anticoagulation or present with severe symptoms, endovascular thrombectomy (EVT) has emerged as a favorable treatment option.

**Objective:** The primary aim of this study is to evaluate the outcomes, complications, and comorbidities associated with EVT in patients with CVT.

**Methods:** A query of the 2015-2019 National (Nationwide) Inpatient Sample (NIS) was performed for patients admitted to hospitals with ICD-10 diagnosis codes for CVT. The usage of endovascular thrombectomy was identified using ICD-10 procedure codes. Demographic information, baseline comorbidities, complications, and discharge dispositions were compared between patients who underwent EVT and those who were managed medically. A multivariate logistic regression was performed to determine independent predictors of outcomes, while controlling for age, gender, cerebral edema/herniation, hemorrhage, mechanical ventilation, Elixhauser Comorbidity Index, and National Institute of Health Stroke Scale (NIHSS) severity. Odds ratios (OR) were calculated for each complication/outcome. Statistical significance was set at an alpha level of 0.05. All statistical analyses were performed using Statistical Product and Service Solutions (SPSS) Statistical Software Version 29.

**Results:** A total of 36,005 patients diagnosed with CVT were identified from 2015(Q4)-2019; of those, 325 (0.9%) underwent EVT. Patients who underwent EVT were older (49.28±19.08years vs 43.62±23.29 years, p < 0.001), more likely to be female (61.5% vs. 54.8%, p = 0.016), and had higher rates of comorbidities, such as diabetes mellitus (18.5% vs. 13.5%, p = 0.012), hypertension (47.7% vs. 31.3%, p < 0.001), and obesity (13.8% vs. 8.1%, p < 0.001). These patients also presented with more severe neurological symptoms, including higher NIHSS scores (13.15 vs. 6.17, p < 0.001), coma (30.8% vs. 11.2%, p < 0.001), and cerebral edema (43.1% vs. 15.5%, p < 0.001). Patients who instead received medical management had higher rates of sepsis (4.7% vs 1.5%, p = 0.008), and were more likely to undergo subsequent decompressive craniectomy (1.9% vs 0%, p = 0.011).

Patients undergoing EVT had a higher incidence of in-hospital mortality (15.4% vs. 4.6%, OR 1.627, p = 0.007), were less likely to be discharged routinely (29.2% vs. 57%, OR 0.599, p < 0.001), and more likely to be transferred to a skilled nursing facility (44.6% vs. 20%, OR 3.22, p < 0.001). They also experienced higher rates of concomitant conditions, such as pulmonary embolism (7.7% vs. 3%, p < 0.001) and hemiplegia (33.8% vs. 10.5%, p < 0.001).

**Conclusions:** In this nationally-representative analysis, patients with CVT who underwent EVT were older, had higher income, were more likely to be female, and presented with more severe neurological conditions. Our results indicate that EVT in patients with CVT is associated with significant risks, including higher rates of inpatient mortality. Although our analysis attempts to adjust for confounding differences, it remains challenging to fully account for the increased baseline morbidity in patients undergoing EVT. As a result, the poorer outcomes observed in this cohort potentially reflect the severity of illness in these patients rather than the risks associated with EVT itself. Nonetheless, EVT remains an important treatment option for patients who fail medical management. Careful patient selection and tailored management strategies are essential to minimize risks and improve outcomes in this high-risk population. Further studies should focus on developing precise patient selection criteria to better identify which patients with CVT are most likely to benefit from EVT.

## Introduction

Cerebral venous thrombosis (CVT) involves the formation of blood clots in dural venous sinuses and cerebral veins. Standard of care for CVT is medical management with low molecular weight heparin followed by transition to oral vitamin K antagonists. Despite reductions in mortality with medical therapy, death and disability from CVT is 10% to 15%.^1-5^ For patients who fail medical therapy, require rescue therapy, or experience clinical decompensation, endovascular thrombectomy (EVT) has emerged as a potential treatment option.^6^

EVT in CVT enables recanalization and restoration of venous blood flow, which can improve clinical outcomes and reduce the risk of further brain injury. While patients with CVT are at risk of hemorrhagic transformation due to venous infarction, the standard preferred treatment in cases of CVT, with or without hemorrhage, remains medical management with anticoagulation.^6,7^ EVT is considered in cases where patients fail medical therapy or experience clinical decompensation. A previous study using the National Inpatient Sample (NIS) Database evaluating trends in the utilization and outcomes of mechanical thrombectomy for CVT found an increase in utilization between 2005 and 2018, with annual increases of 0.13%.^8^ Current studies do not fully stratify patient comorbid conditions and associated outcomes.^9-12^ A better understanding of the risks and benefits associated with EVT in CVT could refine patient selection and improve post-procedural outcomes.

Complications associated with EVT in CVT include new or expanding hemorrhage, catheter-related complications, including venous perforation, catheter tip fracture, groin hematoma or retroperitoneal hematoma, and recurrent CVT.^6^ Although catheter-related complications were found to be as low as 3% in a recent meta-analysis of EVT for CVT, they can still contribute to poor outcomes.^6^ Therefore, establishing preferred catheter techniques, thorough risk assessment, careful patient selection, and diligent post-procedural care are essential to minimize complications and enhance patient outcomes.

The purpose of this study is to use the NIS Database to determine the number of patients admitted with CVT, how many of these patients undergo mechanical thrombectomy, and to evaluate the effectiveness, outcomes, comorbidities, and complications in patients who receive mechanical thrombectomy compared to other interventions. By analyzing data from 2015 to 2019, this study aims to provide a comprehensive understanding of the effectiveness of mechanical thrombectomy in CVT patients and identify the factors associated with successful clinical outcomes.

## Methods

### Data Source and Patient Selection

The NIS Database, developed by the Agency for Health Care Research and Quality as part of the Health Care Cost and Utilization Program (HCUP), is the largest, publicly-available, all-payer, inpatient care database in the United States. The NIS contains data on approximately seven million hospital stays each year, and includes data from hospitals that account for 97% of all discharges in the country. For the years 2015 through 2019, this sample includes data covering about 35 million inpatient visits. The NIS employs a multistage clustering design to develop a stratified probability sample, representing approximately 20% of all inpatient discharges. The data variables include demographic characteristics, hospital and regional information, inpatient diagnoses and procedures, medications, and discharge disposition. Since the NIS is publicly-available and contains no identifiable information, approval by an institutional review board was not required for this study.

A query of the database was performed to identify hospitalizations for patients admitted with CVT who underwent EVT from 2015 (Q4) to the end of 2019. These patients were identified using diagnostic codes from the *International Classification of Diseases, Tenth Revision, Clinical Modification* (ICD-10-CM) and the *International Classification of Diseases, Tenth Revision, Clinical Modification Procedure Coding System* (Table S1). The data start date of 2015 (Q4) was chosen because the *International Classification of Diseases, Ninth Revision, Clinical Modification* was retired before this time in favor of the *Tenth Revision*.

### Data Characteristics and Outcomes Measured

Demographic characteristics, including age, sex, insurance status, race, and socioeconomic status, were described. As defined by the HCUP, the lower 50th percentile median income category reflects those patients below the 50th percentile in income in the United States, allowing the stratification of outcome by wealth of the patient. ICD-10 codes for type 2 diabetes mellitus (DM), hypertension (HTN), hyperlipidemia, smoking, alcohol abuse, substance abuse, obesity, chronic obstructive pulmonary disease (COPD), congestive heart failure (CHF), anticoagulation/antiplatelet treatment were used to analyze baseline demographics of our cohort. ICD-10 diagnosis codes for cerebral edema, hemiplegia, herniation, coma, mechanical ventilation, Elixhauser Comorbidity Index, and NIHSS were used to evaluate stroke severity. Complications of sepsis, acute kidney injury, deep vein thrombosis (DVT), pulmonary embolism (PE), tracheostomy, aphasia, seizure, and non-traumatic hemorrhage were compared using ICD-10 diagnosis codes. Outcomes of length of stay, discharge disposition (Routine discharge is defined by HCUP as discharge home or self-care), inpatient mortality, and total hospital charges were compared.

### Statistical Analysis

Descriptive univariate statistics were used to evaluate baseline clinical characteristics, comorbidities, complications, and outcomes between the EVT and non-EVT groups. Dichotomous variables were assessed using chi-square tests and presented as number and percentage (%). Continuous variables were analyzed using t-tests and reported as mean and standard deviation (SD). A multivariate logistic regression was performed to determine independent predictors of outcomes, while controlling for age, gender, cerebral edema/herniation, hemorrhage, mechanical ventilation, Elixhauser Comorbidity Index, and NIHSS severity. Odds ratios (OR) with 95% confidence interval (CI) were calculated for each complication/outcome. Statistical significance was set at an alpha level of 0.05. All statistical analyses were performed using SPSSv29.

## Results

A total of 36,005 patients diagnosed with cerebral venous thrombosis (CVT) were identified from 2015 (Q4) -2019. Of this total number of patients, 325 (0.9%) underwent endovascular thrombectomy (EVT) and 35,680 (99.1%) did not.

### Baseline Characteristics and Comorbidities

Patients who underwent EVT were on average older than patients who did not undergo thrombectomy (49.28 ± 19.078 years vs. 43.62 ± 23.294 years, p < 0.001) (Table 1). Of the entire cohort, 54.9% were female and 60.4% were white. While there was no statistically significant difference in the rate of EVT based on race, women were more likely to undergo EVT (61.5% vs. 54.8%, p = 0.016) (Table 1).

**Table 1.** Baseline Demographics and Comorbidities of Patients with Cerebral Venous Thrombosis Undergoing Endovascular Thrombectomy vs. Without Endovascular ThrombectomyOR; Odds Ratio, CI; Confidence Interval, SD; Standard Deviation.

| Variables | Total Cohort<br>(36005) | No Endovascular Thrombectomy<br>(35680, 99.1%) | Endovascular Thrombectomy<br>(325, 0.9%) | Endovascular Thrombectomy |  |
| --- | --- | --- | --- | --- | --- |
|  |  |  |  | OR (CI 95%) | P value |
| Demographics |  |  |  |  |  |
| Female | 19755<br>(54.9%) | 19555 (54.8%) | 200 (61.5%) | 1.319 (1.054-1.651) | 0.016 |
| White Race | 21750<br>(60.4%) | 21540 (60.4%) | 210 (64.6%) | 1.199 (0.954-1.506) | 0.124 |
| Lower 50<sup>th</sup> Percentile Median Income | 17970<br>(49.9%) | 17855 (50%) | 115 (35.4%) | 0.547 (0.435-0.687) | < 0.001 |
| Federally Assisted Insurance | 17715<br>(49.2%) | 17555 (49.2%) | 160 (49.23%) | 1.001 (0.805 – 1.246) | 1 |
| Private Insurance | 14960<br>(41.5%) | 14825 (41.5%) | 135 (41.5%) | 1 (0.801-1.248) | 1 |
| Comorbidities |  |  |  |  |  |
| Diabetes Mellitus | 4890<br>(13.6%) | 4830 (13.5%) | 60 (18.5%) | 1.446 (1.091-1.917) | 0.012 |
| Hypertension | 11305<br>(31.4%) | 11150 (31.3%) | 155 (47.7%) | 2.006 (1.612-2.497) | < 0.001 |
| Hyperlipidemia | 6245<br>(17.3%) | 6170 (17.3%) | 75 (23.1%) | 1.435 (1.107-1.86) | 0.007 |
| Obesity | 2930<br>(8.1%) | 2885 (8.1%) | 45 (13.8%) | 1.827 (1.33-2.509) | < 0.001 |
| Chronic Obstructive Pulmonary Disease | 1765<br>(4.9%) | 1740 (4.9%) | 25 (7.7%) | 1.625 (1.078-2.451) | 0.022 |
| **Congestive Heart Failure** | 395 (1.1%) | 395 (1.1%) | 0 (0%) | 0.989 (0.988-0.99) | 0.055 |
| **Smoker Status** | 4710 (13.1%) | 4645 (13%) | 65 (20%) | 1.67 (1.271-2.196) | < **0.001** |
| **Alcohol Abuse Disorder** | 1805 (5%) | 1790 (5%) | 15 (4.6%) | 0.916 (0.544-1.541) | 0.8 |
| **Chronic Anticoagulant/Antip latelet Use** | 6070 (16.9%) | 6050 (17%) | 20 (6.2%) | 0.321 (0.204-0.505) | < **0.001** |
| **Variables** | **Total Cohort Mean (± SD)** | **No Endovascular Thrombectomy Mean (± SD)** | **Endovascular Thrombectomy Mean (± SD)** | **P Value** |  |
| **Age** | 43.67 (±23.265) | 43.62 (±23.294) | 49.28 (±19.078) | < **0.001** |  |
| **Elixhauser Comorbidity Index** | 3.6896 (±2.20452) | 3.6797 (±2.20205) | 4.7846 (±2.20474) | < **0.001** |  |
| **NIHSS** | 6.4638 (±8.08097) | 6.1667 (±7.92227) | 13.15 (±8.746) | < **0.001** |  |

Although no difference was observed based on insurance status, patients from the lower 50th percentile of income were less likely to undergo EVT (35.4% vs. 50%, p < 0.001) (Table 1). This finding highlights the potential influence of socioeconomic factors on access to advanced interventions, such as EVT, which may impact overall treatment outcomes.

Examination of comorbidities between the EVT and non-EVT subsets revealed no significant differences between the two groups with respect to CHF and alcohol abuse disorder. However, patients undergoing EVT more often had coexisting comorbidities, including DM (18.5% vs. 13.5%, p = 0.012), HTN (47.7% vs. 31.3%, p < 0.001), hyperlipidemia (23.1% vs. 17.3%, p = 0.007), obesity (13.8% vs. 8.1%, p < 0.001), COPD (7.7% vs. 4.9%, p = 0.022), smoker status (20% vs. 13%, p < 0.001), and were less likely to have chronic anticoagulant/antiplatelet use (6.2% vs. 17%, p < 0.001) (Table 1).

Patients who underwent EVT also presented with more severe neurological symptoms, including higher NIHSS scores (13.15 vs. 6.17, p < 0.001) and higher Elixhauser Comorbidity Index (4.8746 vs. 3.6797, p < 0.001) (Table 1).

The EVT group demonstrated significantly higher rates of association with DVT (13.8% vs. 4.6%, p < 0.001), PE (7.7% vs. 3%, p < 0.001), aphasia (18.5% vs. 6.9%, p < 0.001), hemiplegia (33.8% vs. 10.5%, p < 0.001), and seizure (27.7% vs. 20.6%, p = 0.002), compared to their non-EVT counterparts (Table 2). They were also more likely to require tracheostomy (10.8% vs. 2.8%, p < 0.001) (Table 2). However, patients who did not receive thrombectomy suffered from higher rates of sepsis over the course of hospitalization (1.5% vs. 4.7%, p = 0.008) (Table 2).

**Table 2.** Complications During Hospitalization of Patients with Cerebral Venous Thrombosis Undergoing Endovascular Thrombectomy vs. Without Endovascular Thrombectomy OR; Odds Ratio, CI; Confidence Interval.

| Variables | Total Cohort<br>(36005) | No<br>Endovascular<br>Thrombectomy<br>(35680, 99.1%) | Endovascular<br>Thrombectomy<br>(325, 0.9%) | Endovascular<br>Thrombectomy |  |
| --- | --- | --- | --- | --- | --- |
|  |  |  |  | OR (CI 95%) | P value |
| Sepsis | 1695 (4.7%) | 1690 (4.7%) | 5 (1.5%) | 0.314 (0.13-0.761) | **0.008** |
| Deep Vein<br>Thrombosis | 1670 (4.6%) | 1625 (4.6%) | 45 (13.8%) | 3.368 (2.449-4.632) | **< 0.001** |
| Pulmonary<br>Embolism | 1090 (3%) | 1065 (3%) | 25 (7.7%) | 2.709 (1.793-4.092) | **< 0.001** |
| Acute Kidney<br>Injury | 3590 (10%) | 3560 (10%) | 30 (9.2%) | 0.918 (0.629-1.338) | 0.711 |
| Tracheostomy | 1050 (2.9%) | 1015 (2.8%) | 35 (10.8%) | 4.122 (2.887-5.886) | **< 0.001** |
| Aphasia | 2535 (7%) | 2475 (6.9%) | 60 (18.5%) | 3.038 (2.289-4.032) | **< 0.001** |
| Hemiplegia | 3855 (10.7%) | 3745 (10.5%) | 110 (33.8%) | 4.363 (3.459-5.503) | **< 0.001** |
| Herniation | 2450 (6.8%) | 2375 (6.7%) | 75 (23.1%) | 4.207 (3.239-5.464) | **< 0.001** |
| Coma | 4090 (11.4%) | 3990 (11.2%) | 100 (30.8%) | 3.53 (2.783-4.478) | **< 0.001** |
| Mechanical<br>Ventilation | 3755 (10.4%) | 3690 (10.3%) | 65 (20%) | 2.167 (1.648-2.85) | **< 0.001** |
| **Seizure** | 7430 (20.6%) | 7340 (20.6%) | 90 (27.7%) | 1.479 (1.158-1.888) | **0.002** |
| **Cerebral Edema** | 5665 (15.7%) | 5525 (15.5%) | 140 (43.1%) | 4.13 (3.31-5.154) | **< 0.001** |
| **Decompressive**<br>**Hemicraniectomy** | 670 (1.9%) | 670 (1.9%) | 0 (0%) | 0.981 (0.98-0.983) | **0.011** |
| OR; Odds Ratio, CI; Confidence Interval |  |  |  |  |  |

### Outcomes

Given the emerging role of EVT in managing CVT, it is essential to thoroughly examine the clinical outcomes associated with this intervention compared to traditional non-EVT management. Understanding these outcomes provides valuable insights into the efficacy, safety, and overall impact of EVT on patient care. Patients undergoing EVT had, on average, a longer length of hospital stay (13.34 vs 10.24 days, p = 0.001), were less likely to be discharged routinely (29.2% vs. 57%, p < 0.001), less likely to be discharged to home health care (4.6% vs. 12%, p < 0.001), and more likely to be transferred to a skilled nursing facility (44.6% vs. 20%, p < 0.001) (Table 3). Those in the non-EVT group were more likely to undergo decompressive hemicraniectomy (1.9% vs. 0%, p = 0.011) (Table 2). Assessing total costs for both groups, patients in the EVT group had significantly higher total charges ($338,090.06 vs. $146,824.40, p < 0.001), reflecting the increased complexity and resources required for their treatment (Table 3). These higher costs may also exacerbate the financial burden on both healthcare systems and patients, particularly those from lower socioeconomic backgrounds who may already face barriers to access.

**Table 3.**
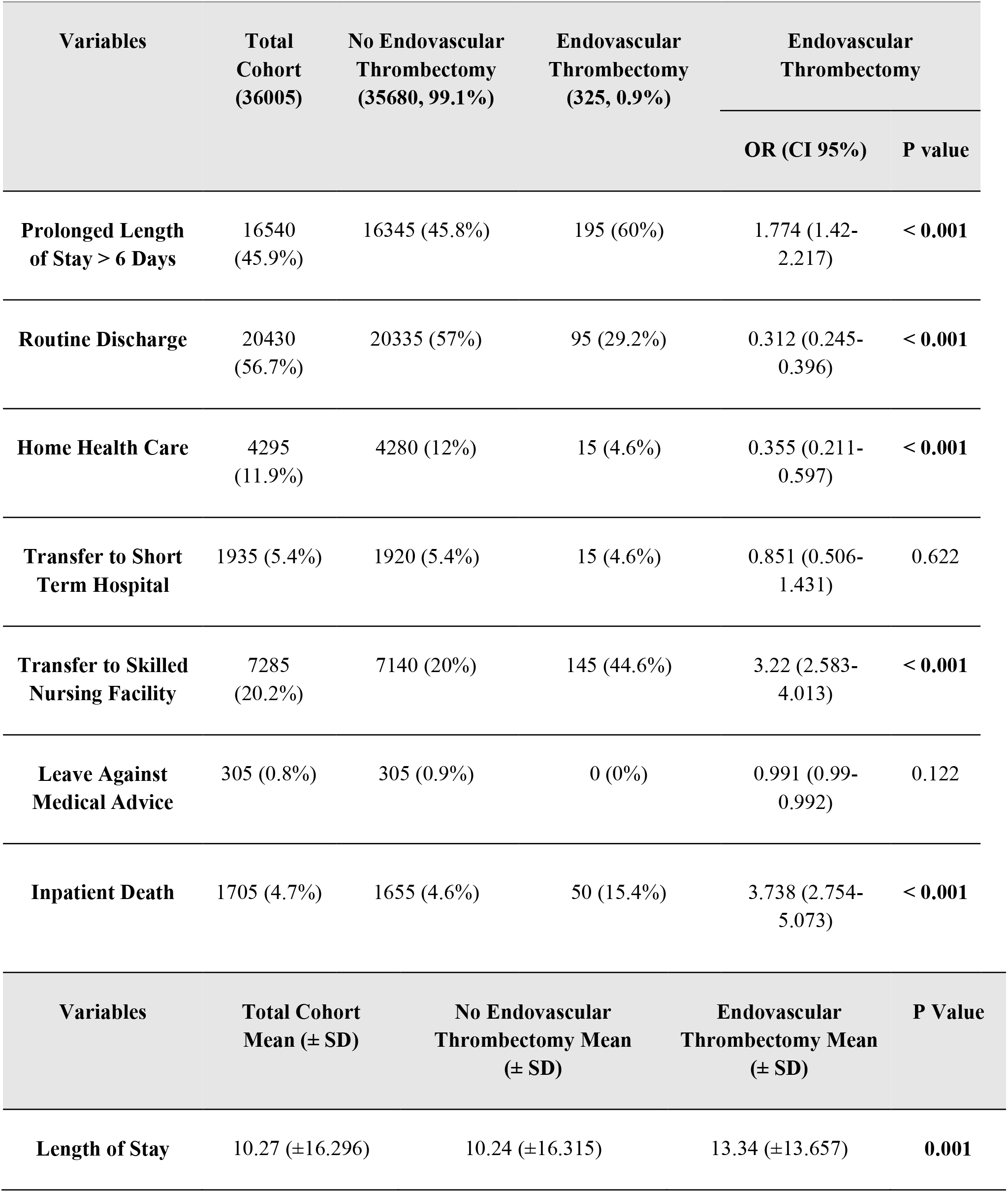
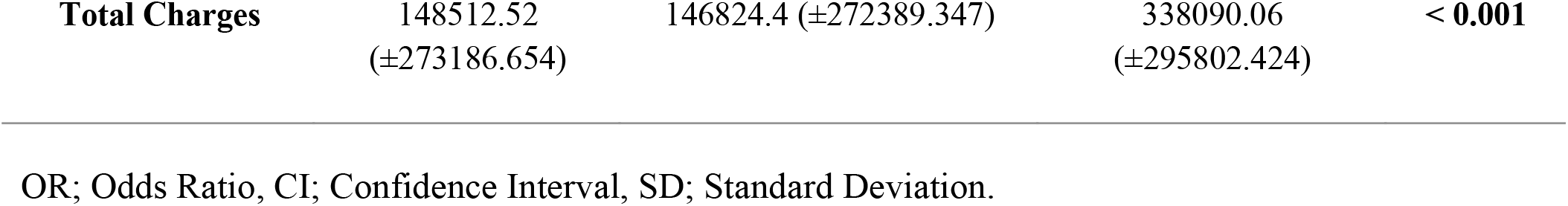
Hospital Course of Patients with Cerebral Venous Thrombosis Undergoing Endovascular Thrombectomy vs. Without Endovascular Thrombectomy OR; Odds Ratio, CI; Confidence Interval, SD; Standard Deviation.

| Variables | Total Cohort<br>(36005) | No Endovascular<br>Thrombectomy<br>(35680, 99.1%) | Endovascular<br>Thrombectomy<br>(325, 0.9%) | Endovascular<br>Thrombectomy |  |
| --- | --- | --- | --- | --- | --- |
|  |  |  |  | OR (CI 95%) | P value |
| **Prolonged Length of Stay > 6 Days** | 16540 (45.9%) | 16345 (45.8%) | 195 (60%) | 1.774 (1.42-2.217) | < **0.001** |
| **Routine Discharge** | 20430 (56.7%) | 20335 (57%) | 95 (29.2%) | 0.312 (0.245-0.396) | < **0.001** |
| **Home Health Care** | 4295 (11.9%) | 4280 (12%) | 15 (4.6%) | 0.355 (0.211-0.597) | < **0.001** |
| **Transfer to Short Term Hospital** | 1935 (5.4%) | 1920 (5.4%) | 15 (4.6%) | 0.851 (0.506-1.431) | 0.622 |
| **Transfer to Skilled Nursing Facility** | 7285 (20.2%) | 7140 (20%) | 145 (44.6%) | 3.22 (2.583-4.013) | < **0.001** |
| **Leave Against Medical Advice** | 305 (0.8%) | 305 (0.9%) | 0 (0%) | 0.991 (0.99-0.992) | 0.122 |
| **Inpatient Death** | 1705 (4.7%) | 1655 (4.6%) | 50 (15.4%) | 3.738 (2.754-5.073) | < **0.001** |
| Variables | Total Cohort<br>Mean (± SD) | No Endovascular<br>Thrombectomy Mean<br>(± SD) | Endovascular<br>Thrombectomy Mean<br>(± SD) | P Value | |
| **Length of Stay** | 10.27 (±16.296) | 10.24 (±16.315) | 13.34 (±13.657) | **0.001** | |

|  |  |  |  |  |
| --- | --- | --- | --- | --- |
| **Total Charges** | 148512.52<br>(±273186.654) | 146824.4 (±272389.347) | 338090.06<br>(±295802.424) | **< 0.001** |

In order to eliminate the severity of neurological condition as a confounding factor, a multivariate logistic regression controlling for age, gender, Elixhauser Comorbidity Index, NIHSS, coma, mechanical ventilation, cerebral edema, and herniation was performed to determine the independent predictors of outcome in our dataset. From this, it was determined that undergoing EVT was positively correlated with inpatient death (OR: 1.627, CI: 1.142-2.317, p = 0.007) and non-routine discharge (OR: 0.599, CI: 0.454-0.79, p < 0.001), even when the previously stated factors are controlled for (Table 4).

**Table 4.** Independent Predictors of Outcomes in Patients with Cerebral Venous Thrombosis Undergoing Endovascular Thrombectomy OR; Odds Ratio, CI; Confidence Interval, SD.

| Variable | Inpatient Death |  | Routine Discharge |  | Prolong Length of Stay > 6 Days |  |
| --- | --- | --- | --- | --- | --- | --- |
|  | OR (CI 95%) | P value | OR (CI 95%) | P value | OR (CI 95%) | P value |
| Endovascular Thrombectomy | 1.627 (1.142-2.317) | 0.007 | 0.599 (0.454-0.79) | <.001 | 1.12 (0.873-1.437) | 0.372 |

## Discussion

This study utilized data from the NIS between 2015 and 2019 to evaluate the baseline characteristics, comorbidities, complications, and outcomes associated with EVT treatment in patients with CVT. We evaluate and discuss our findings in comparison to existing systematic reviews and the TO-ACT trial to help better understand how patient comorbidities and baseline characteristics may correlate with complications and outcomes in CVT patients undergoing EVT versus medical management.^13^

Our study assessed baseline characteristics, including age, gender, and NIHSS on presentation. Results revealed that patients undergoing EVT were generally older, more likely to be female, and presented with more severe neurological symptoms when compared to patients in the non-EVT group. Patients receiving standard medical therapy were more likely to be on chronic anticoagulation and antiplatelet therapy compared to the EVT group. These findings are consistent with the existing literature indicating that patients selected for EVT often have more severe neurological presentations and are more likely to experience advanced disease manifestations, such as encephalopathy or coma at stroke onset.^14-16^

Interestingly, our study identified a higher prevalence of certain comorbidities among the EVT group, including DM, HTN, hyperlipidemia, obesity, COPD, and a history of smoking. There was no significant difference in rates of acute kidney injury, CHF, or chronic alcohol abuse between both cohorts. The significantly higher rates of comorbid conditions found in patients undergoing EVT in CVT compared to the non-EVT group is not previously reported in literature and may contribute to the complexity of patient management, indicating the importance of individualized treatment approaches. Furthermore, patients from lower-income groups were less likely to undergo EVT, suggesting that socioeconomic barriers may influence access to advanced therapies. This highlights the need for addressing disparities in care to ensure equitable access to potentially life-saving interventions.

### Outcomes of EVT

The EVT group demonstrated higher rates of DVT, PE, aphasia, and seizures, compared to the non-EVT group. These findings are consistent with prior studies, where seizures and other complications were more common in patients undergoing EVT.^7,17^ Conversely, patients who did not undergo EVT were more likely to develop sepsis and require decompressive hemicraniectomy. This emphasizes the critical need for meticulous patient selection and risk stratification to minimize procedural risks and optimize outcomes. The TO-ACT trial observed a higher incidence of ICH in the non-EVT group compared to the EVT group.^13^ However, the results were not statistically significant, reflecting the scarce evidence with regards to EVT complications.

### Impact on Clinical Outcomes and Prognosis

Our results show that patients undergoing EVT have worse prognostic factors, including longer hospital stays, a lower likelihood of routine discharge or discharge home, a higher likelihood of transfer to skilled nursing facilities, and a higher rate of inpatient death compared to those in the non-EVT group. The mortality rate observed in our study (15.4%) aligns with the higher end of reported rates in other studies, which range from 3% to 12% depending on patient selection and disease severity. ^10, 14, 18^ These outcomes emphasize the complex post-procedural course of EVT in CVT patients, suggesting that while EVT may offer benefits in terms of recanalization and potential for improved outcomes in selected cases, it also carries significant morbidity and mortality risks.

### Clinical Implications and Future Directions

Our findings indicate that EVT should be considered a viable treatment option for select CVT patients who fail anticoagulation therapy and have no contraindications to surgical intervention. The significantly higher comorbid burden and severe presentations observed in EVT patients convey the need for individualized treatment strategies that consider the unique risk profiles and clinical conditions of each patient. Future studies should focus on refining patient selection criteria, optimizing EVT techniques, and exploring the use of EVT in less severe CVT cases to improve overall outcomes. Additionally, more granular data on procedural techniques and operator experience could help identify best practices and minimize complications, thereby improving patient outcomes.

## Limitations

The primary limitations of this study stem from the use of a large national administrative database, which include a non-randomized retrospective design, potential coding errors, and selection biases. While multivariate logistic regression was used to control for confounders, such as age, NIHSS, and comorbidities, residual confounding related to unmeasured variables -such as clinical decision-making and the timing of interventions -remains. Furthermore, the lack of granular clinical data limits the ability to establish causality. An example of this is the lack of clarity with information extrapolated from NIS as to whether the concomitant conditions were truly concomitant in nature or occurred as post-procedure complications. Nevertheless, our study leverages nationwide data, which enhances the generalizability and external validity of our findings. This broader scope reduces the biases associated with single-center studies, thereby providing a more comprehensive assessment of the impact of EVT therapy on outcomes in CVT.

## Conclusions

Our study shows that patients undergoing EVT for CVT typically have greater baseline comorbidities and more severe presentations, contributing to the observed higher rates of complications and poorer prognostic factors compared to non-EVT patients. While EVT itself may carry risks, the outcomes are potentially reflective of the underlying severity of illness in this group. Therefore, EVT should be considered as an essential option for patients who fail medical management, with careful patient selection and management strategies needed to mitigate risks and improve outcomes. Future research should focus on developing more precise patient selection criteria by incorporating predictive biomarkers, advanced neuroimaging techniques, and risk stratification models. Additionally, exploring the efficacy of EVT in less severe CVT cases, alongside innovations in catheter technology, may reduce complication rates and expand the use of EVT. Finally, studies on long-term functional outcomes following EVT, particularly in different socioeconomic groups, will help guide clinical decisions and improve the overall management of CVT.

## Data Availability

This study data can be requested from the corresponding author after completing the required procedures outlined by the Healthcare Cost and Utilization Project.

## Declarations

### Ethical approval

Since the NIS is publicly-available and contains no identifiable information, approval by an institutional review board was not required for this study.

## Data Tables

## Supplementary Data

**Supplemental Table S1.** Diagnosis and Procedure Codes.

| Diagnosis or Procedure | International Classification of Diseases, 10th Revision, Clinical Modification and Procedure Codes |
| --- | --- |
| Cerebral Venous Sinus Thrombosis | I63.6, I67.6, O22.5, O87.3, G08 |
| Endovascular Thrombectomy | 03CG, 03CH, 03CJ, 03CK, 03CL, 03CM, 03CN, 03CP, 03CQ, 05CL3ZZ |
| Acute Kidney Injury | N17, N28.9 |
| Alcohol Abuse | F10 |
| Anxiety | F41.9 |
| Aphasia | R47.01 |
| Autistic Disorder | F84.0 |
| Bipolar Disorder | F30 |
| Cerebral Edema | G93.6 |
| Cerebral Herniation | G93.5 |
| Chronic Obstructive Pulmonary Disease | J40, J41, J42, J43, J44 |
| Chronic Renal Failure | N18 |
| **Coma** | **R40.2** |
| **Congestive Heart Failure** | **I50.9** |
| **Deep Vein Thrombosis** | **I82.4, I82.6** |
| **Depression** | **F32** |
| **Epilepsy** | **G40** |
| **Hemiplegia** | **G81** |
| **Hemorrhage** | **I60, I61, I62** |
| **Hyperlipidemia** | **E78.5** |
| **Hypertension** | **I10** |
| **Mechanical Ventilation** | **5A0935Z** |
| **Morbid Obesity** | **E66.2** |
| **Obesity** | **E66.9** |
| **Peripheral Vascular Disease** | **I73** |
| **Pulmonary Embolism** | **I26.9** |
| **Schizophrenia** | **F20** |
| **Seizures** | **R56** |
| **Sepsis** | **A41.9** |
| **Smoker** | **Z72.0, F17.2** |
| **Substance Abuse** | **F19.1** |
| **Tracheostomy** | **F024GCZ, 5A1935Z, 5A945Z, 5A1955Z, 0B11** |
| **Type 2 Diabetes Mellitus** | **E11** |

